# A Voting Ensemble Approach for Hepatitis Disease Detection

**DOI:** 10.1101/2020.08.02.20166850

**Authors:** Shawni Dutta, Samir Kumar Bandyopadhyay

## Abstract

The inflammation of the liver is termed as Hepatitis. Several different types of hepatitis are from A to G. For example, Hepatitis A is caused by the hepatitis A virus. Similarly, other type of Hepatitis virus is formed by the name, say Hepatitis G. Some types of virus will not create any serious problems. LongAlasting and cause scarring of the liver, loss of liver function and in some cases, liver cancer are also caused by this disease. Voting ensemble based approach is proposed in this paper as final phase classification that accepts top two classifier models obtained from first and second phase classification respectively. The reason of using the proposed classifier is to enhance the prediction performance so that patients with hepatitis disease are identified correctly.

## 1. Introduction

Hepatitis causes inflammation of liver. The symptoms are swelling, potentially resulting in permanent damage of liver. The disease is quite painful and its effects make the victims feel weak. These infections can be acute or chronic and people can even die from these infections. It is caused by many different factors such as infectious organisms, chemical toxins, poisons, drugs, and alcohol. Viral hepatitis is caused by one of six different types of viruses: hepatitis A, hepatitis B, hepatitis C, hepatitis D, hepatitis E, or hepatitis G. Medical diagnosis is a challenging task which requires timely and accurate identification. Liver is one of the vital parts of our body parts. Presence of hepatitis virus in liver can damage can hamper patients’ lives. According to (1), around 1.34 million deaths occurred due to this disease in 2015. The World Health Organization (WHO) has fixed target of eliminating hepatitis B, hepatitis C virus within 2030.

For accompanying the process of medical diagnosis process, an intelligent model can be suggested. Machine Learning (ML), a subfield of artificial intelligence (AI), applies statistical methods on sample data in order to achieve the best performance result. ML algorithms have been paying substantial attention in health domain in recent years. ML is capable of processing huge dataset and analysis that data in order to provide clinical insights. This will of course assist medical experts in providing utmost care incorporating optimised expenditure. Use of ML in health analytics is necessary since it can accelerate the rate of patients’ satisfaction. This research approaches to build predictive model that includes ML algorithms in order to learn underlying relationships among the data and later can make predictions from those data (2). The purpose of this study is to develop an automated tool that can provide insights to hepatitis affected patients. Life expectancy detection of a patient is really a challenging task which is covered in this paper. In this context, a predictive model is advised that will use supervised ML based classification techniques. The classification technique separates hepatitis affected patients from normal patients. HighestAefficiency and minimized error vulnerable model is favoured in medical data analysis process.

The working paradigm of this research proceeds through multiple steps of classification. During the first step of classification, various algorithms such as Support Vector Machine (SVM) (3), naïve Bayes (NB) (4), kANearest neighbor (KANN) (5), and Decision tree (DT) (6) are implemented. AdaBoost (7), Gradient Boost (GB) (8), and Random Forest (RF) (9) are ensemble based classifications which are implemented as phaseA2 classifiers. All these supervised classifiers will be applied on Hepatitis database and predictions will be evaluated with respect to some preAdefined metrics. After evaluation, the best two predictive models (one from phase 1 and one from phase 2) are picked up and given as input for next step of classification. In this step, voting ensemble classifier is proposed that assembles the predictions of top two models retrieved from first step of classification. The target of the proposed classifier is to attain maximized prediction performance with correct result and lowest error rate.

## 2. Related Works

Hepatitis is a liver disease of variable in nature. Patients with persistent infection by HBV are often associated with chronic liver disease. It leads to the development of cirrhosis and hepatocellular carcinoma (HCC). Researchers also suggest that HBV is not directly cytopathic for the infected hepatocyte (10A12). It is a serious global public health problem. Presently six distinct types of hepatitis virus are identified and called as hepatitis A, B, C, D, E and G viruses. The primary source of infection is the faeces with fecalAoral route being the most predominant mode of transmission for Hepatitis A virus (HAV) and Hepatitis E virus (HEV) (13A16). Blood borne viruses are Hepatitis B virus (HBV), Hepatitis C virus (HCV), and Hepatitis D virus (HDV) (17A18). It is primarily transmitted through a breach in the skin (percutaneous) or mucosa (mucosal). Hepatitis viral infections are acute but hepatitis B, C and Delta can result in chronic infections.

Several researches have been carried out in the field of hepatitis disease detection. For obtaining predictive tool for hepatitis disease diagnosis, SVM, KANN, Artificial Neural Network (ANN) algorithms are implemented. A comparative study is drawn among these classifiers in order to obtain the best predictive module (19). Another study (20) implemented naïve bayes, J48, RF, and MLP for recognizing hepatitis patients. A combined research has been proposed in (21) that hybridizes support vector machine (SVM) and simulated annealing (SA). 10Afold crossAvalidation method is used for estimating the classification procedure. Another research (22) used NonAlinear Iterative Partial Least Squares to accomplish the data dimensionality reduction, SelfAOrganizing Map technique for clustering performance and ensembles of NeuroAFuzzy Inference System for predicting the hepatitis disease. Advantage of ensemble technique used in (22) has shown effective hepatitis diagnosis prediction.

## 3. Dataset Used

This research applies classification methods on real world dataset retrieved from kaggle machine learning repository (23). The dataset consists of number of records and each record is formulated as a collection of several attributes. Table 1 depicts summary of collected dataset. In this dataset, attribute ‘class’ defines the live or dying tendency of patients. This attribute is kept as dependent variable or target variable for classification. Figure 1 shows the distribution of target variable on the dataset. Once the dataset is obtained, it is partitioned into training and testing dataset with the ratio of 67:33. Data classification proceeds in two phasesAtraining phase and testing phase. The training dataset is used to train the model during the training phase. Later the trained model is used for disease classification and the life expectancy of the Hepatitis person.

**Table 1:** Summary of the collected dataset.

| Attributes | Values |
| --- | --- |
| Class | Die (1), Live (2) |
| Age | 7-78 |
| Sex | male(1), female(2) |
| Steroid | no(1), yes(2) |
| Liver Big | no(1), yes(2) |
| Liver Firm | no(1), yes(2) |
| Fatigue | no(1), yes(2) |
| Malaise | no(1), yes(2) |
| anorexia | no(1), yes(2) |
| Spiders | no(1), yes(2) |
| Ascites | no(1), yes(2) |
| Antivirals | no(1), yes(2) |
| Bilirubin | 0.3-8.0 |
| Varices | no(1), yes(2) |
| Spleen Palpable | no(1), yes(2) |
| Alkaline Phosphate | 26-295 |
| serum glutamic-oxaloacetic transaminase (SGOT) | 14-648 |
| Albumin | 2.1-6.4 |
| Pro-time | 0-100 |
| Histology | no(1), yes(2) |

**Figure 1:**
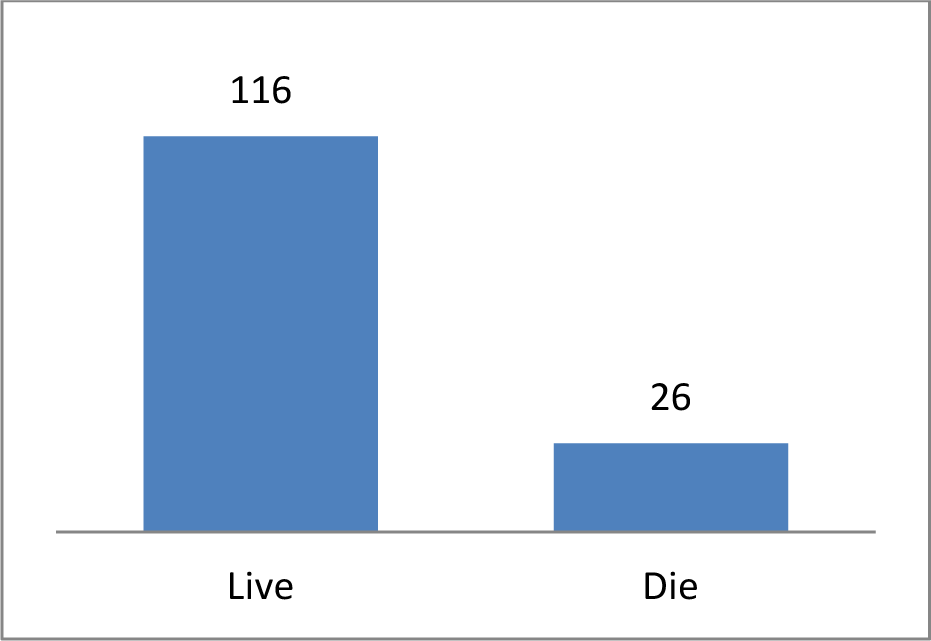
Life expectancy distribution in the dataset

## 4. Methodology

The objective of this research is to provide insight to hepatitis affected patients so that utmost care can be offered to them. For this purpose, an intelligent predictive model is approached that accepts interfering factors which cause hepatitis. The intelligent automated tool should be efficient enough to correctly identify patients with lowest error rate. Several supervised ML algorithms are utilized in this context. The proposed methodology proceeds through 3 steps which are explained as in this section and diagrammatic description is provided in Figure 2.

**Figure 2.**
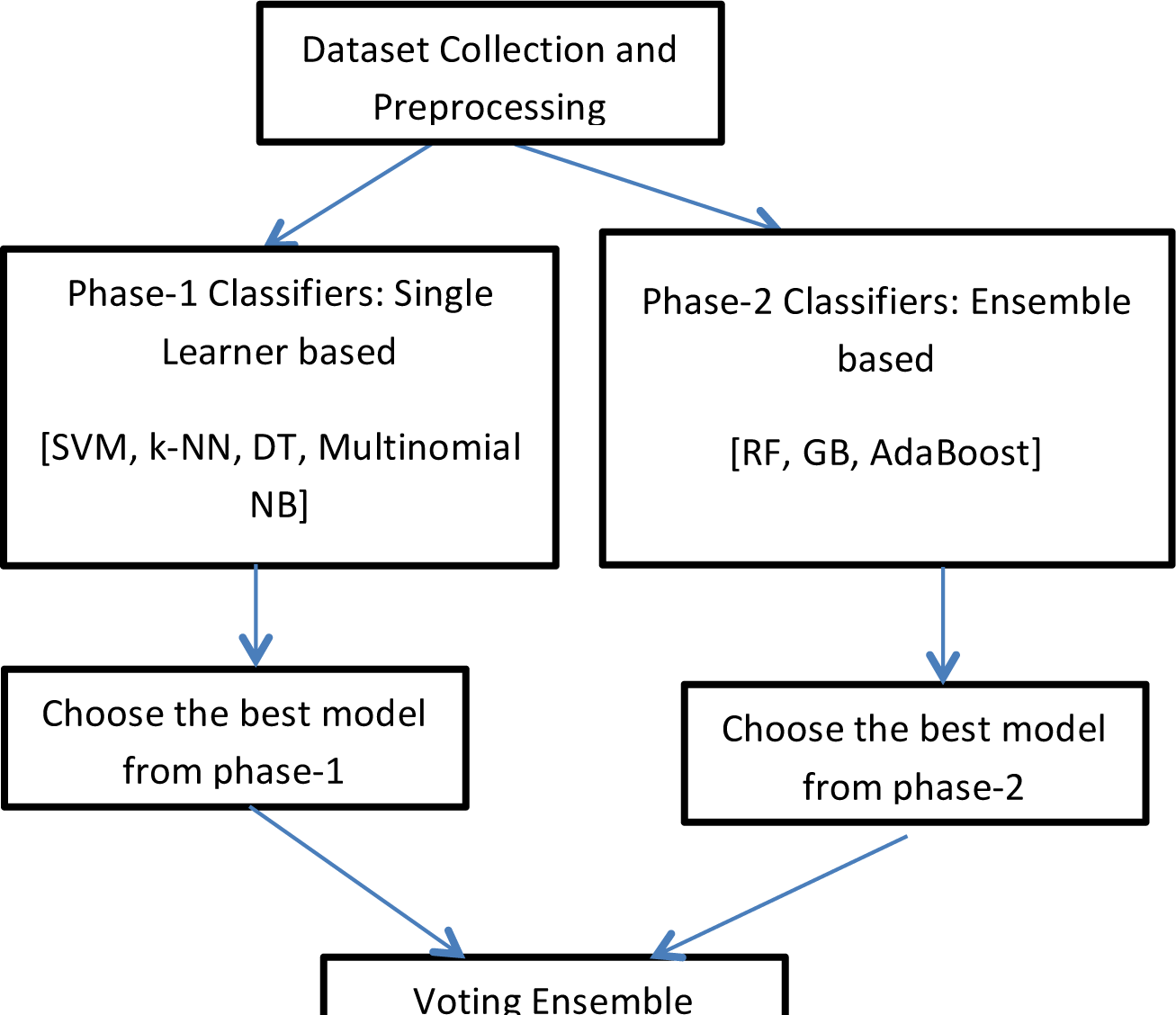
Proposed Methodology

- During the first phase/step, single learner based classifier model is implemented. Decision Tree (DT), KANearest Neighbor (KANN), Support Vector Machine (SVM), Naïve Bayes (NB) classifier models are implemented as phaseA1 classifications.
- In the next step, ensemble based classifiers such as Random Forest (RF), Gradient Boosting (GB) and AdaBoost are applied.
- From each of the above mentioned phases, the best model is selected based on certain evaluation process. These two models are given as input to voting ensemble based classifier model. The target of this proposed voting based strategy is to enhance the prediction result.

## 5. Background

The proposed predictive model is based on supervised ML algorithms. Classification techniques, supervised ML algorithms, solve the problem of assigning class labels to the instances of specified problem domain. In other words, these techniques are applied to dataset for predicting the class labels for unknown cases. Generally, classification techniques exemplify the use of training dataset so that example instances of the problem domain can be mapped efficiently to class labels. This section provides describes details of all the classification techniques those are implemented in phase 1 and phase 2 while designing the planned framework.

### 5.1 PhaseP1 Classification algorithms

The Naïve Bayes (NB) classifier is a statistical model that corresponds to simplest form of Bayesian network. A Bayesian network is considered to be as acyclic directed graphs where nodes present in the graph denotes variables and the links among the nodes represent casual influence among the variables. This classifier exemplifies the use of conditional independence assumption which is infrequently correct in most of the realAworld applications. By demonstrating the use of statistical method along with supervised technique this method obtains classification result (14). NB classifier provides promising results in practice even if the assumed estimates are inaccurate. The accuracy of this classifier is not related to feature dependencies rather than it is the amount of information loss of the class due to the independence assumption is needed to predict the accuracy (4).

KANearest Neighbour Classifiers (5), are simple and effective nonAparametric classification approach. It is often known as lazy learners that identify objects based on closest proximity of training examples in the feature space. While classifying an instance m, its k nearest neighbours are identified which in turn form neighbourhood of instance m. However, the main challenge of this classification technique relies on deciding the appropriate value of k.

Support Vector Machine (SVM) (3) can handle classification tasks with superior generalization performance. It can map input vector to a higher dimensional space by constructing a maximal separating hyperAplane. Basically two parallel hyperAplanes are constructed on each side of the hyperAplane that separates the data. The two parallel hyperA planes are separated by a plane called, separating hyperAplane. It maximizes the distance between the two parallel hyperAplanes. The maximized distance between these parallel hyperAplanes are considered to obtain better generalization error by the classifier [27].

A Decision Tree (DT) (6) exemplifies the use of treeAlike structure. Each goal variable is denoted as a leaf node of DT and nonAleaf nodes of DT are used as a decision node that indicates certain test. The outcomes of those tests are identified by either of the branches of that decision node. Classification results are obtained from this model by starting from the beginning at the root this tree are going through it until a leaf node is reached. This can be useful for forecasting the goal based on some criterion by implementing and training this model (6).

### 5.2 PhaseP2 Classification algorithms

Ensemble techniques are popular ML methods that assemble individually trained set of classifier models. The target of ensemble methods is to produce prediction result which is more accurate and has less error with respect to single learners (25). *Random Forest (RF)* classifier is supervised ensemble machine learning algorithm. This classifier creates decision trees on randomly selected data samples, gets prediction from each tree and decides on the best solution by means of voting (9).

Boosting technique can construct new predictive model while combining the predictions of multiple weak learners. This technique mainly focuses on reducing misclassification rate (25). *AdaBoost* is known to be the first boosting technique proposed by Freund and Schapire. This classifier is known as a metaAestimator that proceeds by fitting a classifier on the original dataset and additional copies of the classifiers are fitted after reAweighting the incorrectly classified instances in such a manner that the classifier is capable in handling more difficult cases (7). *Gradient Boosting (GB)* (8) algorithm is another boosting algorithm that consecutively fits new models for obtaining promising accuracy in estimating the response variable. This allows new baseAlearners to be maximally correlated with the negative gradient of the loss function, associated with the whole ensemble.

### 5.3 Implementation Details

This section gives description of phaseA1 and phaseA2 classifiers about how it is implemented. Parameter tuning is an important task that is concerned of identifying the correct values. Choosing correct parameter values will assist in obtaining the best predictive result. After checking several parameter values, the best parameter value is chosen for corresponding classifier model. Best parameter values used for these classifiers are summarized in table 2.

**Table 2:** Implementation Details of Classifier models.

| Phase-1 Classifiers | Classifier Model | Parameters Used |
| --- | --- | --- |
|  | DT | 1. Criterion: Gini<br>2. Splitter:best |
|  | K-NN | 1. Value of k=7<br>2. Distance Metric=Minkowski |
|  | SVM | 1. Regularization Parameter (c)=1<br>2. Kernel: RBF |
|  | Naïve Bayes | 1. Used Multinomial NB<br>2. Alpha value=1.0 |
| Phase-2 Classifiers | RF | 1. Criterion: Gini<br>2. Number of Base estimators: 100 |
|  | AdaBoost | 1. Number of Base estimators: 500<br>2. Learning Rate:1.0 |
|  | GB | 1. Number of Base estimators: 500<br>2. Learning Rate:1.0<br>3. Loss Function: Deviance<br>4. Split Quality Criterion: Friedman MSE |

#### Voting Ensemble Method

Using voting strategy, it is potential to make a good choice out of multiple possible solutions. Multiple classification techniques may be applied on the same dataset and predictions are acquired. Classifiers actually cast their preference for one or more solutions. Considering majority preferences, final decision is drawn for problemAsolving approach. It is possible to obtain a better solution when several potential algorithms work towards the same problem domain. This voting strategy is advantageous because all of the classifiers will not make the same mistake while instance classification.

For instance classification, voting ensemble method is implemented either using ‘Hard’ or ‘Soft’ voting. In case of hard voting, prediction is made based on choosing majority votes. This means that, the prediction which collects majority votes is selected for final result. However, ascending sort order is utilized when tie occurs. In case of soft voting, prediction probabilities obtained from individual models are summed up and then the prediction having the largest sum is chosen. With soft voting, predictions from some models can be assigned using either uniform or dissimilar weights (26).

In this paper, one best performing model is chosen from phaseA1 classifiers and another model is chosen from phaseA2 classifiers. Finally, the predictions from these individual models are assembled using soft voting ensemble classifier. The prediction obtained from this soft voting ensemble method is retrieved as final result.

### 5.3 Performance Evaluation Metrics

While justifying the efficiency of predictive results, it is necessary to put some concentration of some predefined metrics. A confusion matrix is used to visualize performance of a particular algorithm. In terms of predictive analysis, confusion matrix is considered to be collection of two rows and two columns. False positive (FP), False Negative (FN), True Positive (TP) and True Negative (TN) are reported using this matrix. These terms are explained in terms of disease detection.

TP: If an input is correctly classified as die case by classifier model.

TN: If an input is correctly classified as live case by classifier model.

FP: If an input is classified as die case which is originally a live case.

FN: If an input is classified as live case which is originally a die case.

Using these TP, TN, FP, FN terms, several metrics such as accuracy, precision, recall, f1Ascore can be calculated. Accuracy is a metric that detects the ratio of true predictions over the total number of instances considered (27). It can be defined as equation (1)

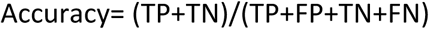

Precision identifies the ratio of correct positive results over the number of positive results predicted by the classifier. Recall denotes the number of correct positive results divided by the number of all relevant samples. F1AScore or FAmeasure is a parameter that is concerned for both recall and precision and it is calculated as the harmonic mean of precision and recall (28). Precision, recall, f1Ascore can be defined as equation (2), (3), (4) respectively.

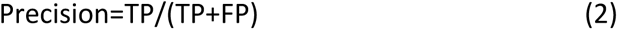

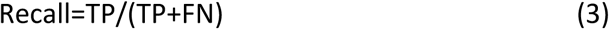

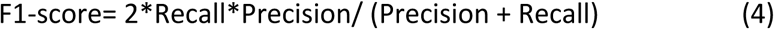

Mean Squared Error (MSE) is another evaluating metric that measures absolute differences between the prediction and actual observation of the test samples (28).

## 6. Experimental Results

This section tries to find out the performance of the best predictive model. From the implemented phaseA1 classifiers, the best model is retrieved based on accuracy, f1Ascore and MSE. After comparing the prediction results, Multinomial NB turns out to be the best classifier model. Again, from phase 2 the best model is selected. RF classifier is the best model from phaseA2. Now, Multinomial NB and RF classifier is fed as input voting ensemble method. This model is capable enough to boost up the prediction results to that of Multinomial NB and RF classifier. The individual models cannot reach much higher accuracy. To obtain enhanced efficiency, soft voting ensemble method is utilized. The proposed voting ensemble method assembles the results of Multinomial NB and RF classifier models and finally reaches an accuracy of 89.36%, f1Ascore of 0.89 and MSE 0.11. All the prediction results are shown in Table 3.

**Table 3:** Performance Comparison of Classifier Models.

|  | **Accuracy** | **F1-Score** | **MSE** |
| --- | --- | --- | --- |
| **Phase-1 Classifier Models** |  |  |  |
| DT | 74.47% | 0.74 | 0.26 |
| K-NN | 82.98% | 0.83 | 0.17 |
| SVM | 82.98% | 0.83 | 0.17 |
| Multinomial NB | 87.23% | 0.87 | 0.13 |
| **Phase-2 Classifier Models** |  |  |  |
| RF | 85.11% | 0.85 | 0.15 |
| GB | 80.85% | 0.81 | 0.19 |
| AdaBoost | 82.98% | 0.83 | 0.17 |
| **Proposed Model** |  |  |  |
| Voting Ensemble | 89.36% | 0.89 | 0.11 |

## Conclusions

Hepatitis is the disease of liver caused by virus. Its widespread impact on human population there is continuous search of new medicine for its treatment and reduces the chance of disease. This study detects the feasibility of using ML approaches while detecting hepatitis disease. Early detection of this disease may assist medical experts to suggest counter measures. The feasibility of using ML techniques is accomplished by implementing an intelligent predictive model. This intelligent model is based on voting ensemble strategy that assembles the prediction of Multinomial NB and RF classifier model those are obtained as two best predictive models over the peer models. The proposed model can recognize hepatitis patients efficiently with minimized error rate. Experimental results have shown that voting ensemble based classifier model can be assessed with promising accuracy of 89.36%, f1Ascore of 0.98 and minimized MSE of 0.11.

## Data Availability

The data is available freely in the Internet.

https://www.kaggle.com/harinir/hepatitis

